# Analyzing Chinese interest in osteoporosis-related diseases by using the Baidu index

**DOI:** 10.1101/2023.11.02.23297966

**Authors:** Jianjun Wu, Yanping Lin, Jingyi Chen, Jiachun Huang, Hongxing Huang

**Affiliations:** The Third School of Clinical Medicine, Guangzhou University of Chinese Medicine, Guangzhou, Guangdong Province, China; The Third Affiliated Hospital of Guangzhou University of Chinese Medicine, Guangzhou, Guangdong Province, China

**Keywords:** osteoporosis, Chinese interest, osteoporosis-related diseases, Baidu index

## Abstract

**Purpose:** Understanding the public’s concern about osteoporosis in China is crucial for guiding public health campaigns and educational efforts, as it is significant public health concern in country. Using the Baidu index, this study aims to examine the Chinese fascination with diseases associated with osteoporosis.

**Methods:** The research gathered information on the search frequency of osteoporosis-related terms on Baidu between January 2012 and December 2022. To gauge the popularity of subjects associated with osteoporosis, the Baidu index was utilized. The study analyzed the trends and patterns of search volume and identified the most popular topics related to osteoporosis.

**Results:** The study found an increasing trend in interest in osteoporosis-related diseases in China over the past ten years. The search volume for osteoporosis-related keywords increased steadily from 2012 to 2022. The most popular diseases related to osteoporosis were hyperosteogeny, fracture, osteoarthritis, and metabolic diseases.

**Conclusions:** The study offers valuable information about the level of understanding and awareness among the general population in China regarding diseases associated with osteoporosis. The growing fascination with subjects related to osteoporosis indicates that the general population is developing a greater understanding of the significance of maintaining bone health. The results of the study can provide valuable guidance for public health campaigns and educational efforts focused on enhancing the prevention and management of osteoporosis-related conditions in China.

---

Osteoporosis (OP) is a condition affecting the entire skeletal system, marked by reduced bone density, deterioration of bone structure, heightened susceptibility to fractures^(1)^. OP can occur at any age but is more common in postmenopausal women and the elderly^(2)^. According to the initial epidemiological investigation on OP in China, it has been observed that individuals aged 50 and above in China are facing a significant health concern due to osteoporosis. OP affects approximately 19.2% of individuals aged 50 and above, particularly middle-aged and elderly women who have a prevalence rate of 32.1%^(3)^. The prevalence of OP among individuals in their middle and old age is concerning as the aging process intensifies^(4)^.

The rise of search engines, specifically, has greatly transformed individuals’ lives, enhancing the Internet’s value as a resource for daily activities, education, and career opportunities. According to the 51st Statistical Report on Internet Development in China, as of December 2022, the number of Internet users in China reached approximately 1.067 billion, resulting in an Internet penetration rate of 75.6%^(5)^. It is estimated that about 81.3% of Internet users use search engines. By using this service, a total of 77.3% of people can obtain the information they require. Baidu Search, which was utilized by 90.9% of users ^[6]^, emerged as the top choice among search engines in China.

The Baidu Index is a platform that allows the sharing of data derived from Baidu’s extensive records of Internet user behavior. It enables the analysis of search trends for keywords, provides valuable insights into shifts in user demands, tracks trends in public opinion across media, and identifies digital consumer traits. The industry perspective allows for the analysis of market characteristics as well. Currently, researchers have utilized Baidu index big data to examine health data, encompassing diverse subjects such as assessing the demands for nursing services and the primary factors that affect them in urban China^(6)^,as well as evaluating the spatiotemporal trends of public panic levels in China amidst the pandemic crisis^(7)^. By examining these online search patterns, one can observe the search habits and preferences of individuals on the internet, which can provide insights into the health information sought by the public. No research has been conducted on OP using Baidu Index.

In order to understand the characteristics of popular interest in OP, this research utilizes Baidu index data platform to observe online search behavior through Internet search data. It also investigates the significance of Internet search data in tracking trends in media attention. Through this system, the public can better understand OP, prevent, and treat OP, and complement traditional OP monitoring systems.

## Methods

### Data source

The data was acquired using Baidu Index. This data sharing platform uses data from the Baidu search engine to share user behavior information. One can study keyword search trends, gain insight into shifts in user requirements, monitor trends in public opinion in the media, and analyze the characteristics of digital consumers to understand their positioning. Additionally, market characteristics can be examined from an industry standpoint.

The study employed the subsequent data: (1) Search index: The scientific evaluation and calculation of the weighted search frequency of every keyword in the Baidu online search were conducted using keywords as the statistical subject, based on the search volume data of internet users on Baidu. (2) Information index: news connected to specific keywords that are indexed by Baidu News and published by major online media. (3) Yearly search rate on the Internet: search index / total number of netizens (from the total number of annual users in the current year of China’s Internet Development Statistical Report).

The search term ‘osteoporosis’ was queried on the Baidu Index platform’s demand map. The most popular OP-related terms were chosen, including “hyperosteogeny,” “fracture,” “osteoarthritis,” and “metabolic diseases.” excluding words that refer to diseases that are not related to osteoporosis. Collected the information index for each keyword spanning from 2018 to 2022, along with the search index for every keyword ranging from 2012 to 2022.

### Statistical methods

We combined the keyword indexes searched for each year to create a single yearly search index. This index was then utilized to examine the characteristics and trends of media and public interest in OP. A one-way ANOVA was conducted for each keyword to compare the yearly, quarterly, and annual Information indexes. Additionally, we explored the correlation between the index and the year using the Pearson correlation method. The statistical difference in the correlation line angle between the groups was determined by conducting analysis of covariance after creating a scatterplot and calculating the correlation coefficient for the users’ search rate each year. P<0.05 (two tailed) defines statistical significance. The figures were created using Apple numbers (Apple, Inc., Cupertino, CA, USA) and GraphPad Prism version 9.5.1 (GraphPad software, Inc., Boston, Ma, USA), and all statistical analyses were performed using GraphPad Prism.

## Results

### Changes to search index

From 2012 to 2019, the annual search index has consistently increased for every keyword, indicating a positive correlation with the corresponding year (Pearson’s correlation coefficient 0.7942; P = 0.0186). Despite a sharp decline in the annual search index amount for each phrase in 2020 (with an increment rate of −34.18%), the overall annual search index for each keyword has been steadily increasing over time. It has reached a significantly higher level in 2022 compared to 2019 (with an increment rate of 47.14%).

The total annual search index for each keyword has exhibited a steady increase over the past ten years (2012–2022). In Figure 1, there was a strong negative correlation (Pearson’s correlation = −0.8475, P = 0.001) between fracture and the year, whereas the year showed a significant positive correlation (Pearson’s correlation = 0.9589, P = 0.0001) with osteoarthritis. There was no correlation between hyperosteogeny and metabolic diseases (hyperosteogeny: Pearson’s correlation = 0.3862, P = 0.2408; metabolic diseases: Pearson’ s correlation = 0.4015, P = 0.2210). The hyperosteogeny search index experienced the largest growth, with an increase of 1,551,543, followed by osteoarthritis with a growth of 119,419. Hyperosteogeny had the highest growth rate (211%) among the search index, with osteoarthritis following closely behind (97%). However, the fracture search index exhibited a decrease of −42,290, accounting for a decline of 10%. Table 1 displays the progression of every search index over a span of ten years, including the correlation between each alteration and the corresponding years.

**Figure 1.**
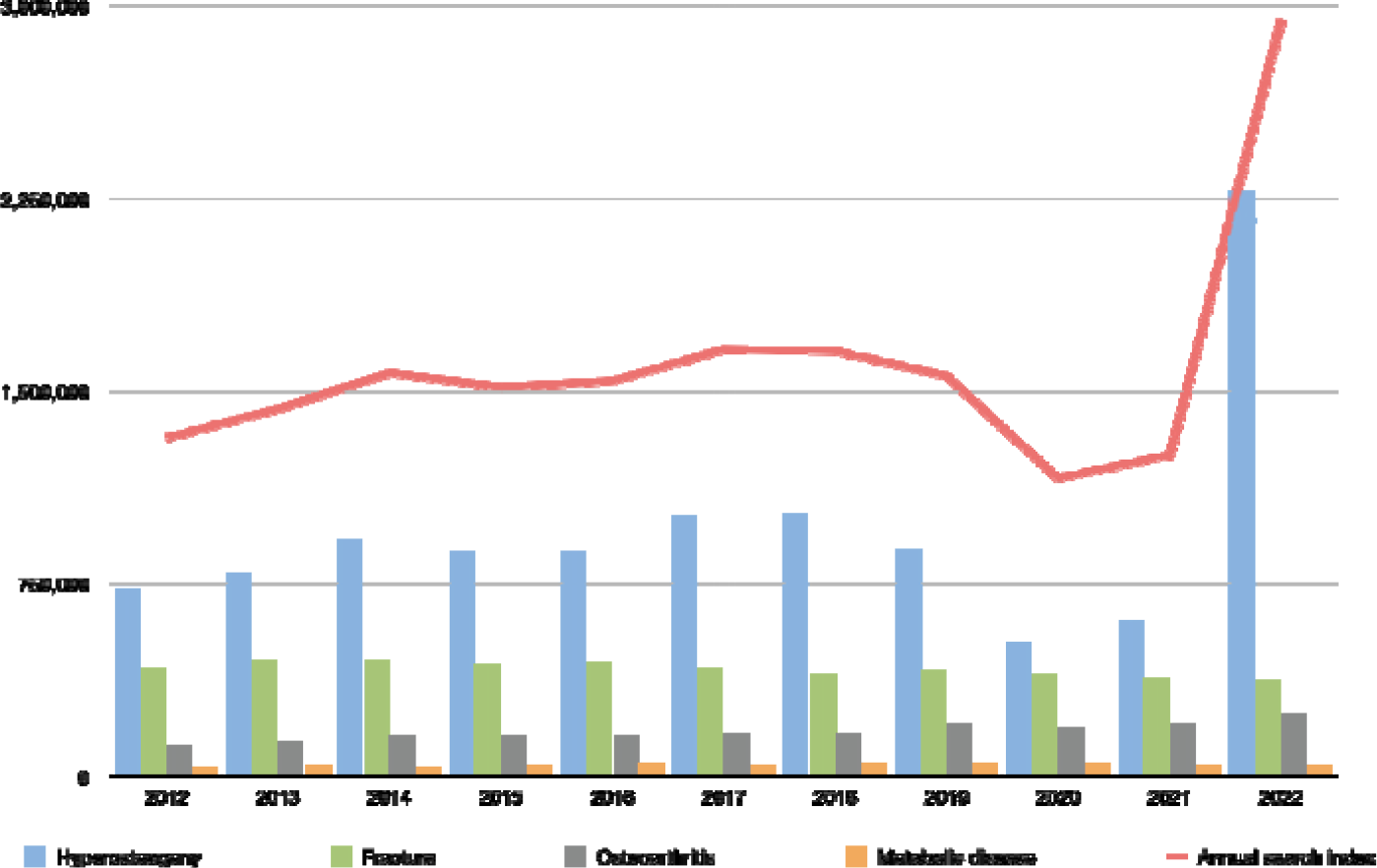
Annual changes to search index.

**Table 1.**
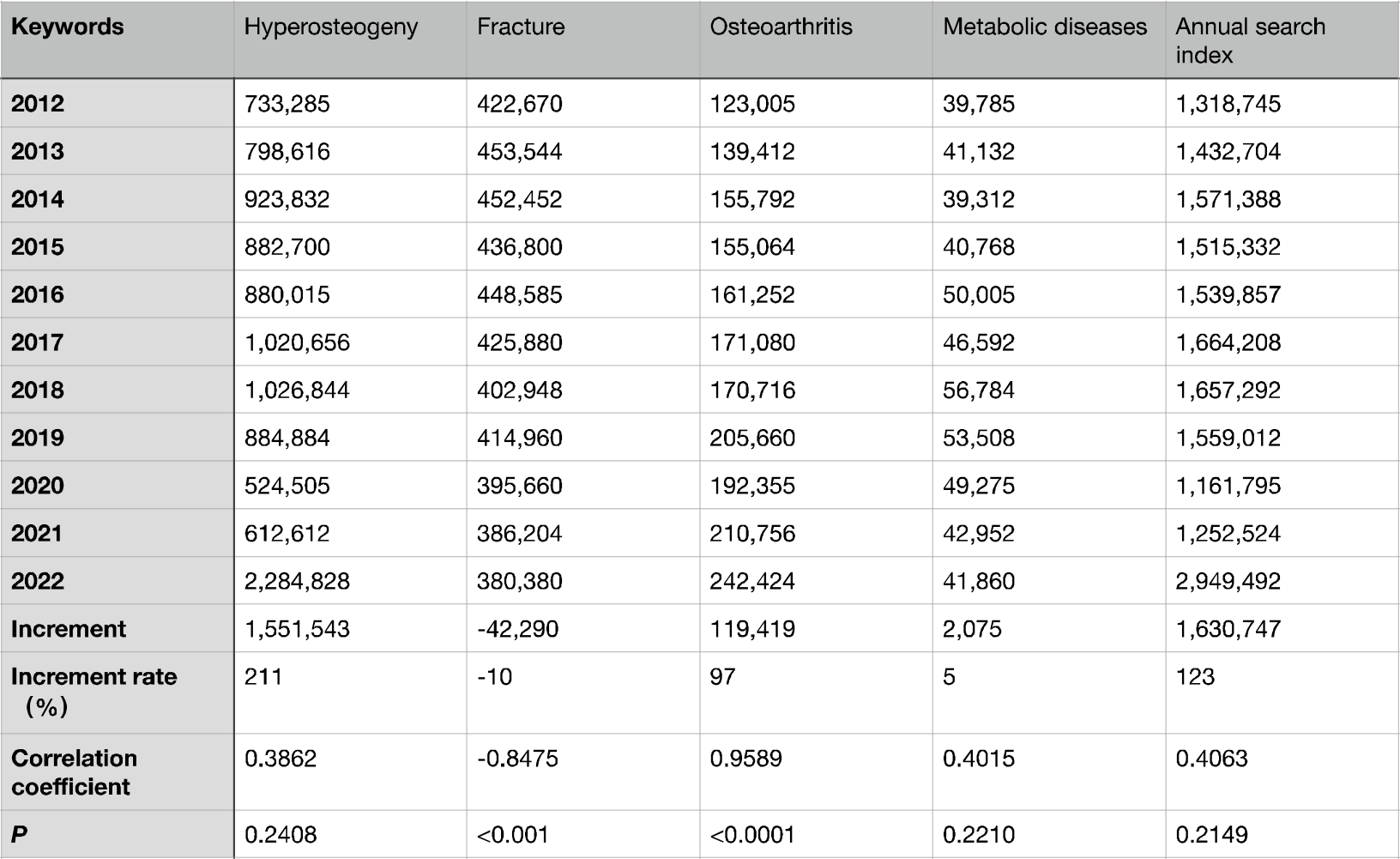
Analyses of the search index’s fundamental status and correlation for the period from 2012 to 2022.

The Fisher’s least significant difference method found a discrepancy in statistics between hyperosteogeny and fracture, hyperosteogeny and osteoarthritis search indexes, hyperosteogeny and metabolic diseases (P <0.0001), and between fracture and metabolic diseases (P = 0.0030). However, fracture and osteoarthritis did not statistically differ from one another (P = 0.0833), osteoarthritis and metabolic diseases (P = 0.5690). In comparison to the other three keywords, hyperosteogeny had the greatest search index during the past ten years, followed by fracture, osteoarthritis and metabolic diseases. Table 2 illustrates the specific results.

**Table 2.** Comparisons from search index keywords.

| Keywords |  | P | 95% CI |  |
| --- | --- | --- | --- | --- |
| Hyperosteogeny | Fracture | <0.0001 | 273739 | 808569 |
|  | Osteoarthritis | <0.0001 | 518517 | 1053348 |
|  | Metabolic disease | <0.0001 | 648112 | 1182943 |
| Fracture | Osteoarthritis | 0.0833 | -22637 | 512194 |
|  | Metabolic disease | 0.0030 | 106958 | 641789 |
| Osteoarthritis | Metabolic disease | 0.5690 | 137821 | 397010 |

According to Figure 2, over the last ten years, apart from the fracture search index, there has been a rise in the yearly search rate of internet users, with all linear regression slopes exceeding 0. Nevertheless, the covariance analyses indicated that there was no statistically significant difference in the slope of the linear regression between the groups. (F = 1.598, P = 0.2067).

**Figure 2.**
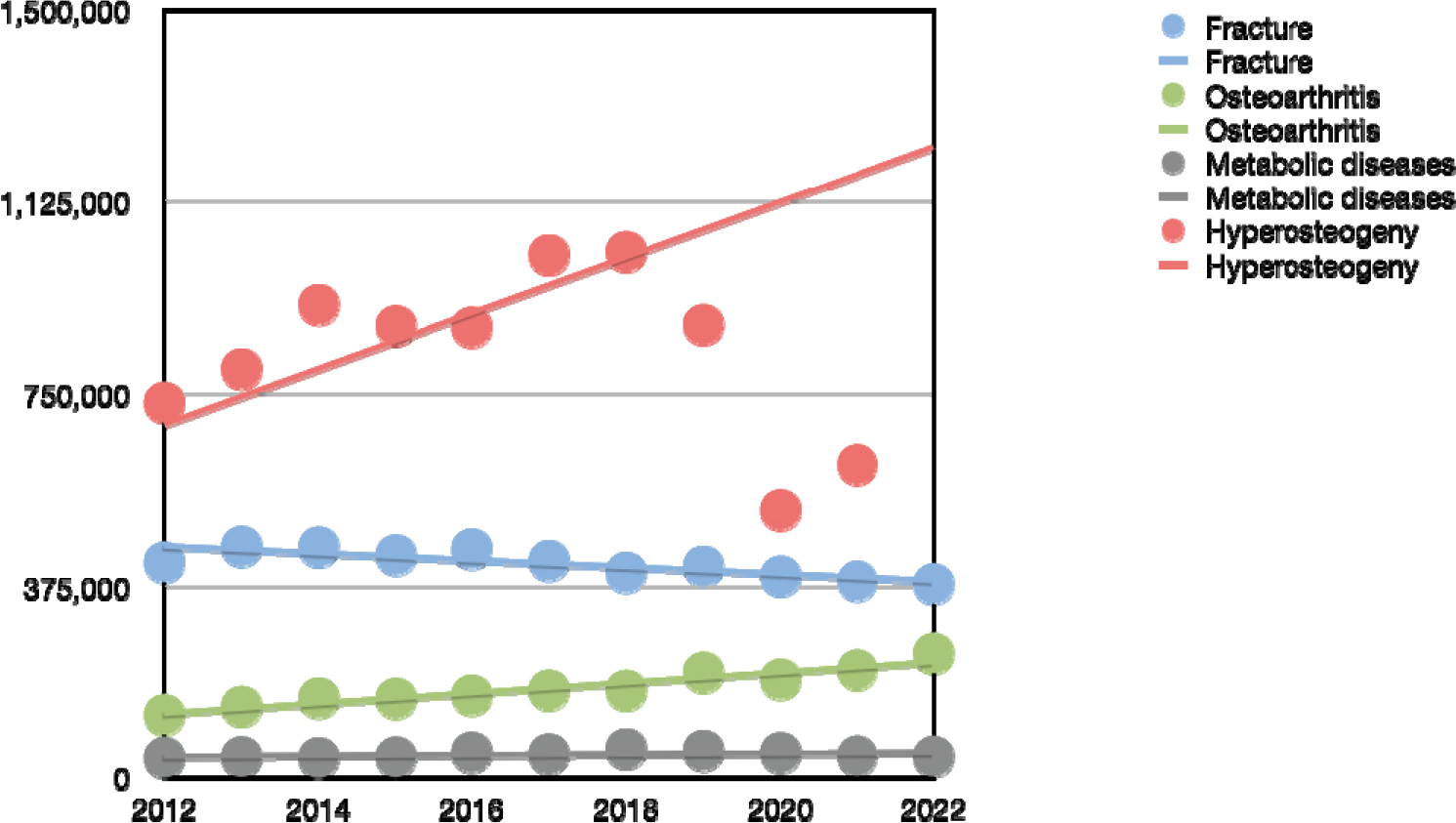
Annual netizen search rate scatter plot.

### Changes to information index

From 2018 to 2022, the information index exhibited a comparable pattern to that of the search index. Table 3 displays the fluctuation in the information index of each keyword over a period of five years. When compared to osteoarthritis and metabolic illnesses (P = 0.0458), the information index of fracture showed significant differences from those conditions (P = 0.0343) (See Supplemental Table 1).

**Table 3.** Basic information index situation from 2018 to 2022.

| Keywords | 2018 | 2019 | 2020 | 2021 | 2022 |
| --- | --- | --- | --- | --- | --- |
| Hyperosteogeny | 17,391 | 76,879 | 70,876 | 41,419 | 43,966 |
| Fracture | 261,952 | 208,384 | 113,447 | 93,617 | 493,838 |
| Osteoarthritis | 7803 | 18,361 | 18,540 | 11,291 | 33,260 |
| Metabolic diseases | 0 | 1 | 0 | 0 | 60 |

## Discussion

During the past decade, the study discovered an increase in public awareness regarding OP; however, the level of attention towards specific diseases varied. Hyperosteogeny and fracture garnered considerably more attention when compared to osteoarthritis and metabolic diseases. Furthermore, the search index for hyperosteogeny achieved a superior ranking compared to the remaining three keywords, surpassing the second place by more than twice the magnitude. This may be connected to the recent rise in the occurrence of hyperosteogeny. Hyperosteogeny and osteoporosis have a strong connection, as the depletion of calcium in bones can result in osteoporosis, while the irregular accumulation of calcium in bones can lead to hyperosteogeny^(8-10)^.

OP is a condition that tends to be hidden, with many individuals not experiencing any symptoms in the beginning stages^(11)^. However, when discomfort arises, patients often take the initiative to gather relevant information through personal research^(12, 13)^. The fact that Baidu is China’s most widely used search engine means that its search results can fairly reflect the needs of its users. The number of confirmed COVID-19 cases in China can be monitored using the illness prediction tool developed collaboratively by Baidu and the Center for Systems Science and Engineering at Johns Hopkins University^(14)^. In addition to conventional detection methods, it can also be utilized to predict the outbreak pattern of diseases^(15)^. The utilization of the Baidu index in OP-related research has not been fully exploited in China. Our research, which is the initial investigation into the behavior and interest of Chinese internet users in OP, validates the potential of utilizing internet search data to depict the real situation of OP patients in China.

Regarding the information index, the findings revealed that fracture received more attention from the media than other keywords. This demonstrates how, during the last five years, the public now has access to more fracture-related information via the media. Overall, the OP was receiving more media coverage. The explanation may be connected to the Internet’s quick development, the population’s aging trend, increased hygiene awareness, the emergence of “Internet Plus Healthcare,” and the rise of OP-related media.

Despite the fact that the population affected by OP incurred significant healthcare costs in China^(16)^, there has consistently been a dearth of information regarding the outbreak of OP in the country. It is extremely challenging to grasp the true prevalence of OP and understand the requirements or characteristics of OP patients. In recent years, there has been research on web-based digital disease monitoring methods due to the rising popularity of Internet usage and the public’s increasing dependence on Internet search engines for obtaining health information. Search engines can disclose sensitive information about diseases before disclosing a diagnosis as a query tool, helping to improve disease control. Big data from the Internet has many potential uses in the medical industry and might be used to enhance and expand on present clinical and observational data^(17-19)^. Given the quick growth of Internet access and engines of search, network analysis of data might prove considered as a supplement to today’s more traditional illness monitoring techniques.

## Limitations

There are a few other restrictions on this study. First of all, we only pay attention to OP attention from users of the Baidu search engine; We neglected to take into account OP attention from users of social media or users of other search engines. This may only represent the concern of the public. Second, the Baidu index could be influenced by sampling bias. Despite the significant increase in China’s Internet penetration rate, there remains a noticeable preference towards socioeconomically and educationally privileged groups among netizens. Thirdly, the study’s China-specific focus, the data had to include a small number of foreigners. Furthermore, assessing the efficiency and reliability of the Baidu Index proves challenging due to the undisclosed details of the method’s particular criteria, despite the implementation of a weighted filtering approach for processing all Baidu Index data. Fourth, due to privacy protection law, Information index data was not made public before 2018. To ensure data variation, future studies should examine data obtained from different engines of search and media. To enhance the objectivity of the research findings, it is imperative to meticulously analyze the data, thereby mitigating the influence of confounding variables.

## Conclusions

From 2012 to 2022, there was a rising trend in the rate of online searches for OP. However, amidst the COVID-19 pandemic, both the search rate and the information index exhibited a decline. By utilizing the Baidu index, individuals can monitor the online behaviors and areas of interest of Chinese Internet users. Improving our understanding of disease prevalence, patient education, and the availability of online resources could be beneficial. Tracking people’s requests for information about OP using data on searching on the Internet trends can be valuable. Furthermore, apart from serving as a scientific basis for the prevention and management of OP in China, it can also be employed as a research method to gain further insights into the characteristics and preferences of the patients.

Between 2012 and 2022, the online search rate for OP showed an increasing trend; however, during COVID-19, it decreased, as well as the information index showed a comparable pattern. Using the Baidu index, one may track Chinese Internet users’ online activities and OP interest. This could enhance our knowledge of illness incidence rates, education of patients, and the accessibility of resources on the Internet. Tracking people’s requests for information about OP using data on searching on the Internet trends can be valuable. In addition to providing a scientific foundation for the prevention and treatment of OP in China, it can be utilized as a research technique to find out more about the traits and interests of the patients.

## Data Availability

The Baidu Index Data Platform provides the information used in this study. (https://index.baidu.com/v2/main/index.html#/help).

https://index.baidu.com/

## Abbreviations

OP: osteoporosis

## Declarations

### Ethics approval and consent to participate

The Baidu Index belong to public databases. Users can download relevant data for free for research and publish relevant articles. Our study is based on open-source data, so there are no ethical issues and other conflicts of interest.

## Consent for publication

Not applicable

## Authors’ contributions

All authors have contributed to the study of concepts and designs. Manuscript writing and data analysis by J.-J.W., Y.-P.L. performed the word revision and formatting adjustments. The remaining authors carried out the paper revision and grammar editing work. Y.-P.L., J.-Y.C., and J.-C.H. provided technical and thesis writing guidance support throughout the process. Corresponding author H.-X.H. reviewed the article and is responsible for communicating with the editor. All authors read and approved the final manuscript.

## Acknowledgements

We appreciated the Baidu Index Platform for providing the data for the manuscript.

