## Supplementary figures and images for "Analyzing Chinese interest in osteoporosis-related diseases by using the Baidu index"

### Supplemental Table 1

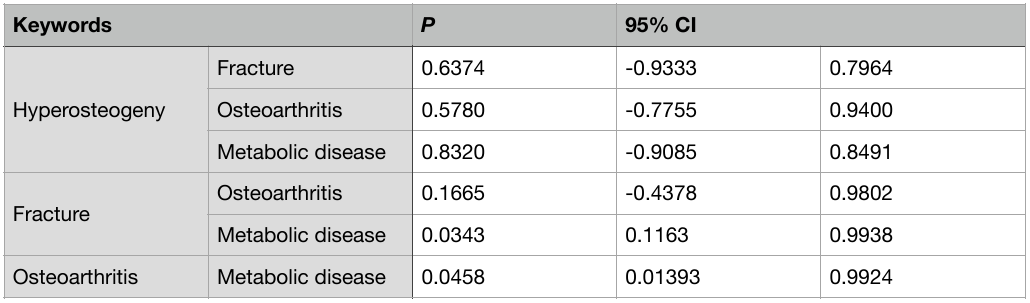

Table 1. Multiple keyword comparisons (Information index).
